# A gender comparison of the psychological distress of medical students in Nigeria during the Coronavirus pandemic: A cross-sectional survey

**DOI:** 10.1101/2020.11.08.20227967

**Authors:** Oluwaseun Mercy Idowu, OyinOluwa Gloria Adaramola, Boluwatife Samson Aderounmu, Ifeoluwa Delight Olugbamigbe, Olaoluwa Ezekiel Dada, Adeyinka Christopher Osifeso, Oluseun Peter Ogunnubi, Oluwakemi Ololade Odukoya

## Abstract

**Background:** The Coronavirus disease (COVID-19) pandemic as a large scale stressor could have negative distress on the mental health on medical students. Since gender differences in mental health may exist between males and females, it would prove interesting to see if a large scale stressor such as the pandemic will cause variances in the psychological distress between both genders.This study assessed and compared the psychological distress of COVID-19 among male and female medical students in medical schools in South-Western Nigeria

**Materials and methods:** A cross-sectional online survey using was carried out among 1010 medical students from three largest universities in south western Nigeria during the COVID-19 pandemic. The respondents were purposively selected, data was obtained on participants demographic and psychological distress was assessed using the General Health Questionnaire 12 (GHQ-12). Data was analyzed using the SPSS version 21 statistical software, chi square was used to assess gender differences, multivariate regression analysis assessed the predictors of psychological distress among both gendersand p values less than 0.05 were considered significant.

**Results:** Female medical students are at a higher risk of psychological distress compared to their male counterparts (p<0.005). Females were almost twice at risk of psychological distress during the COVID-19 pandemic than males (OR=1.534, 95% p=0.003). Females with a positive history of mental illness were five times more at risk of psychological distress during the COVID-19 pandemic compared to females with no previous mental health history (OR=5.102, p=0.002)

**Conclusion:** Females were at higher risk of psychological distress compared to male students. Gender specific interventions addressing psychological distress among medical students are recommended.

## Introduction

The Coronavirus disease 2019 (COVID-19) is caused by the severe acute respiratory syndrome coronavirus-2 (SARS-CoV2), which is highly transmissible and has been a cause for worldwide concern [1]. Since the outbreak of COVID-19 in Wuhan, China in December 2019 [2], the virus has expressed high rates of rapid transmission. It was declared a pandemic on the 11^th^ March, 2020 after more than 118,319 confirmed cases and 4,292 deaths had been recorded from various continents of the world [3]. Shortly before this declaration, Nigeria, the most populous country in Africa, recorded her first case on the 27^th^ February, 2020 [4]. As at the 24th of October 2020, Nigeria had a total of 61,930 confirmed cases with 1,129deaths. Almost half (49%) of these cases are in south-west states of the country [5].

According to the World Health Organization (WHO) mental health is “a state of well-being in which the individual realizes his or her own abilities, can cope with the normal stresses of life, can work productively and is able to make a contribution to his or her community” [6]. A person’s mental health is influenced by social and economic conditions like family, school and social support among other factors [7]. It is widely believed that social ties play a positive part in the preservation of one’s mental health [8]. In a bid to curb the spread of the COVID-19 there has been a disruptionin social life due to the advent of measures like social distancing, lockdown, self-isolation and quarantine. Apart from the social distancing, there is also the possibility of fear and uncertainty in the hearts of many concerning the novel coronavirus which may pose a risk to their mental health [9]. Moreso, several nations have also suffered from economic recessions as a result of the adverse distress of COVID-19 on the economy, leading to loss of employment or reduction in income which could negatively affect mental health [10].

Gender differences in mental health may exist between males and females, with females observed to have a higher prevalence in mental health disorders compared to males. Thismay be because females are exposed to risk factors such as gender inequality, gender-based violence and gender discrimination [11]. A mental health survey carried out in Lagos StateNigeria for the assessment of mental health disorder symptoms, revealed significant gender differences with females showing higher prevalence [12]. Since strategies for identification, prevention and treatment of mental disorders may be based on gender [13], there is a need for a gender specific study when measuring the psychological distress of any population.

Previous studies have shown that the predisposing factors to mental health disorders are gender specific [14-17]. It would prove interesting to see if a large scale stressor such as the pandemic will cause a change in the psychological distress between males and females and help us know if these variances can be characterized as COVID-19 related, hence this study.

This study assessed and compared the psychological distress of COVID-19 among male and female medical students in medical schools in South-Western Nigeria. It also assessed the risk factors associated with psychological distress among male and female medical students. It is hoped that the study findings will be useful for policy makers in the educational sector and provide information useful in tailoring the medical curriculum to suit the peculiar needs of medical students during and after the COVID-19 pandemic.

## Materials and methods

This was a descriptive cross-sectional study thatcompared the psychological distress between male and female medical students of the three largest Colleges of Medicine in the South-western regions of Nigeria. The Colleges are: College of Medicine, University of Ibadan (COMUI), College of Medicine, University of Lagos (CMUL) and Lagos University Teaching Hospital (LASUCOM). Purposive sampling was used to select the three largest medical schools in South-western Nigeria, which the study was carried out. Convenience sampling method was used in recruiting eligible male and femalerespondents (medical and dental undergraduates ofthe three universities), who participated in the study.

### Data collection

This survey was conducted from June 22 to July 16, 2020. This was about 4-5 months after the first case of COVID-19 were reported in Nigeria. Because it was not feasible to do a face-face sampling survey during the on-going pandemic, data was collected using an online survey platform (Google forms https://forms.gle/19yfEzehJKwsme759). Relying on the authors’ networks with colleagues in the three universities, a recruitment poster was created and posted to the class online platform in the three colleges. This poster contained a brief introduction on the background, voluntary nature of participation, declarations of anonymity and confidentiality, as well as the link code of the online questionnaire. Persons, who are medical students of the three colleges, understood the content of the poster and agreed to participate in the study were instructed to complete the questionnaire by clicking the link.

### Survey instrument

The General Health Questionnaire (GHQ-12), a self-administered screening tool was used to assess individuals with psychological distress [18]. It was developed in 1970 by Sir David Goldberg and Paul Williams [19]; the twelve item GHQ-12 is the most extensively used screening instrument for common mental disorders, in addition to being a more general measure of psychiatric well-being. It assesses the severity of mental problems over the past few weeks. Its brevity makes it attractive for use [18,19]. Its psychometric properties have been studied in various countries and in several sub-populations, including in Nigeria [20,21].

#### GHQ scoring

The bimodal scale was used in this study to grade the participants risk of psychological distress during the pandemic, with a score of 0 given to the ‘Not at all’ and ‘No more than usual’ responses while a score of 1 is given to the ‘Same as usual’ and ‘much less than usual’ response. A total summation of the scores between 0 and 12 was collated for the responses selected by each participants, scores ≤ 2 implies that the participants have ‘No risk of psychological distress’, whereas scores ≥ 3 implies that participants are ‘At risk of psychological distress’.

#### Data analysis

Data collected was analyzed with SPSS 21 statistical software for windows (version 21.0 SPSS Inc, Chicago IL). Categorical variables were expressed in frequency tables with the corresponding percentages while normally distributed continuous variables were expressed as means and standard deviations. Chi square was used to assess gender differences in categorical and continuous variables respectively. A multivariate regression analysis was used to identify the predictors of psychological distress first among the general participants, and subsequently for males and females separately. p values of <0.05 were considered statistically significant

##### Ethical considerations

Ethical approval was obtained from Research and Ethics Committee of the Lagos University Teaching Hospital, with HREC assigned number: LUTHHREC/EREV/0620/56. Informed consent (online) was obtained from the participants before the commencement of the study. Participation was voluntary and Confidentiality assured to all respondents. Data was stored anonymously in a password-protected database

## Results

The socio-demographic characteristics of study participants are shown in Table 1. The total number of data collected for male respondents was 486 and that of females, 524.

**Table 1:** Socio-demographics of Male and Female Participants.

| Variables | Male<br>n=486 | Female<br>n=524 | Total | Chi-square<br>p value |
| --- | --- | --- | --- | --- |
|  | Frequency (%) | Frequency (%) |  | (X<sup>2</sup>) |
| **Age group** |  |  |  |  |
| 15-19 | 82 (37.3) | 138 (62.7) | 220 (100) | $X^2 = 35.386$<br>$p < 0.001$ |
| 20-24 | 308 (47.4) | 342 (52.6) | 650 (100) |  |
| 25-29 | 75 (67.0) | 37 (33.0) | 112 (100) |  |
| 30-34 | 18 (72.0) | 7 (28.0) | 25 (100) |  |
| 35-39 | 3 (3.0) | 0 (0.0) | 3 (100) |  |
| *Mean ± SD =* | *22.43±3.221* | *21.28±2.523* |  |  |
| **Ethnicity** |  |  |  |  |
| Yoruba | 362 (48.9) | 379 (51.1) | 741 (100) | $X^2 = 36.645$<br>$p = 0.223$ |
| Igbo | 82 (46.6) | 94 (53.4) | 176 (100) |  |
| Hausa | 3 (3.0) | 0 (0.0) | 3 (100) |  |
| Edo | 12 (42.9) | 16 (57.1) | 28 (100) |  |
| Others | 27 (55.6) | 35 (66.8) | 62 (100) |  |
| **Religion** |  |  |  |  |
| Christianity | 400 (46.6) | 459 (53.4) | 859 (100) | $X^2 = 5.913$<br>$p = 0.052$ |
| Islam | 80 (56.3) | 62 (43.7) | 142 (100) |  |
| Others | 6 (66.7) | 3 (33.3) | 9 (100) |  |
| **Institution** |  |  |  |  |
| COMUI | 169 (56.5) | 130 (43.5) | 299 (100) | $X^2 = 12.028$<br>$p = 0.002$ |
| CMUL | 182 (44.4) | 228 (55.6) | 410 (100) |  |
| LASUCOM | 135 (43.9) | 166 (55.1) | 301 (100) |  |
| **Level** |  |  |  |  |
| Preclinicals | 199 (48.5) | 211 (51.4) | 410 (100) | $X^2 = 4.636$<br>$p = 0.328$ |
| Clinicals | 287 (47.8) | 313 (52.2) | 600 (100) |  |
| **Marital Status** |  |  |  |  |
| Single | 478 (48.3) | 511 (51.7) | 989 (100) | $X^2 = 0.958$<br>$p = 0.619$ |
| Married | 8 (64.3) | 13 (35.7) | 21 (100) |  |
| **Family Monthly income** |  |  |  |  |
| <33,000 | 86 (51.2) | 82 (48.8) | 168 (100) | $X^2 = 3.425$<br>$p = 0.331$ |
| 33,000-50,000 | 75 (52.4) | 68 (47.6) | 143 (100) |  |
| 50,000-100,000 | 97 (49.5) | 99 (50.5) | 196 (100) |  |
| >100,000 | 228 (45.3) | 275 (54.7) | 503 (100) |  |
| **Have you ever been diagnosed of a mental illness** |  |  |  |  |
| Yes | 10 (34.5) | 19 (65.5) | 29 (100) | $X^2 = 2.224$<br>$p = 0.136$ |
| No | 476 (48.5) | 505 (51.5) | 981 (100) |  |
| **Do you have a Relative/Acquaintance with COVID 19** |  |  |  |  |
| Yes | 36 (35.3) | 66 (64.7) | 102 (100) | $X^2 = 7.475$<br>$p = 0.006$ |
| No | 450 (49.6) | 458 (50.4) | 908 (100) |  |

Abbreviations: SD Standard deviation, COMUI College of Medicine, University of Ibadan, CMUL College of Medicine, University of Lagos, LASUCOM Lagos State University Teaching Hospital.

The mean age of female respondents was 21.28 ± 2.5 years and made up more than half (54.7%) of respondents who had a family monthly income greater than 100,000 naira. More females (65.5%) constitute the group of respondents who reported to have been previously diagnosed with a mental condition and also made up 64.7% of respondents who had a relative/acquaintance diagnosed of COVID-19. In comparison, the mean age of males was *22*.*43±3*.*2* years which were higherthan the mean age of females. Male respondents constitute less than half (45.3%) of respondents who had a family income greater than 100,000 naira. Fewer male (34.5%) constituted the group of respondents’ whoreported having ever been diagnosed of a mental illness and males made up 35.3% of respondents who had relatives/acquaintances diagnosed of COVID-19 as compared to females who made up majority (65.5% and 64.7% respectively) of both groups.

### Psychological distress assessment

From the results in Table 2, more females reported they had *lost much sleep over worry (53*.*9%), been feeling depressed (56*.*2%)* and *been thinking of themselves as worthless (54*.*1%)*, ‘Rather more than usual’, than the male gender (46.1%, 43.8% and 45.9% respectively).

**Table 2:**
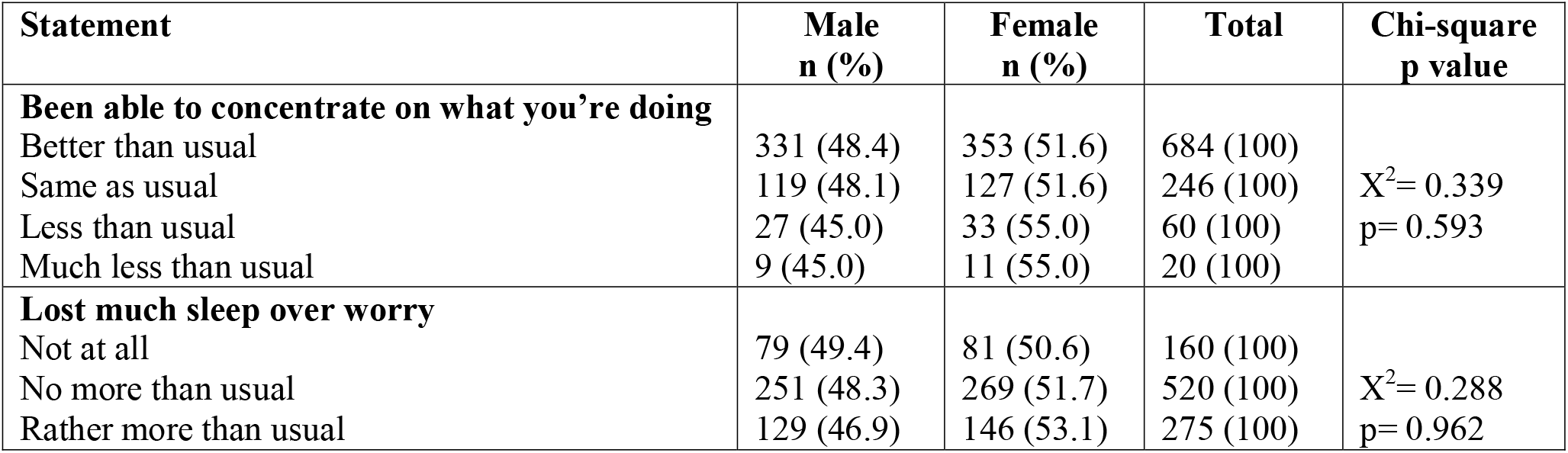
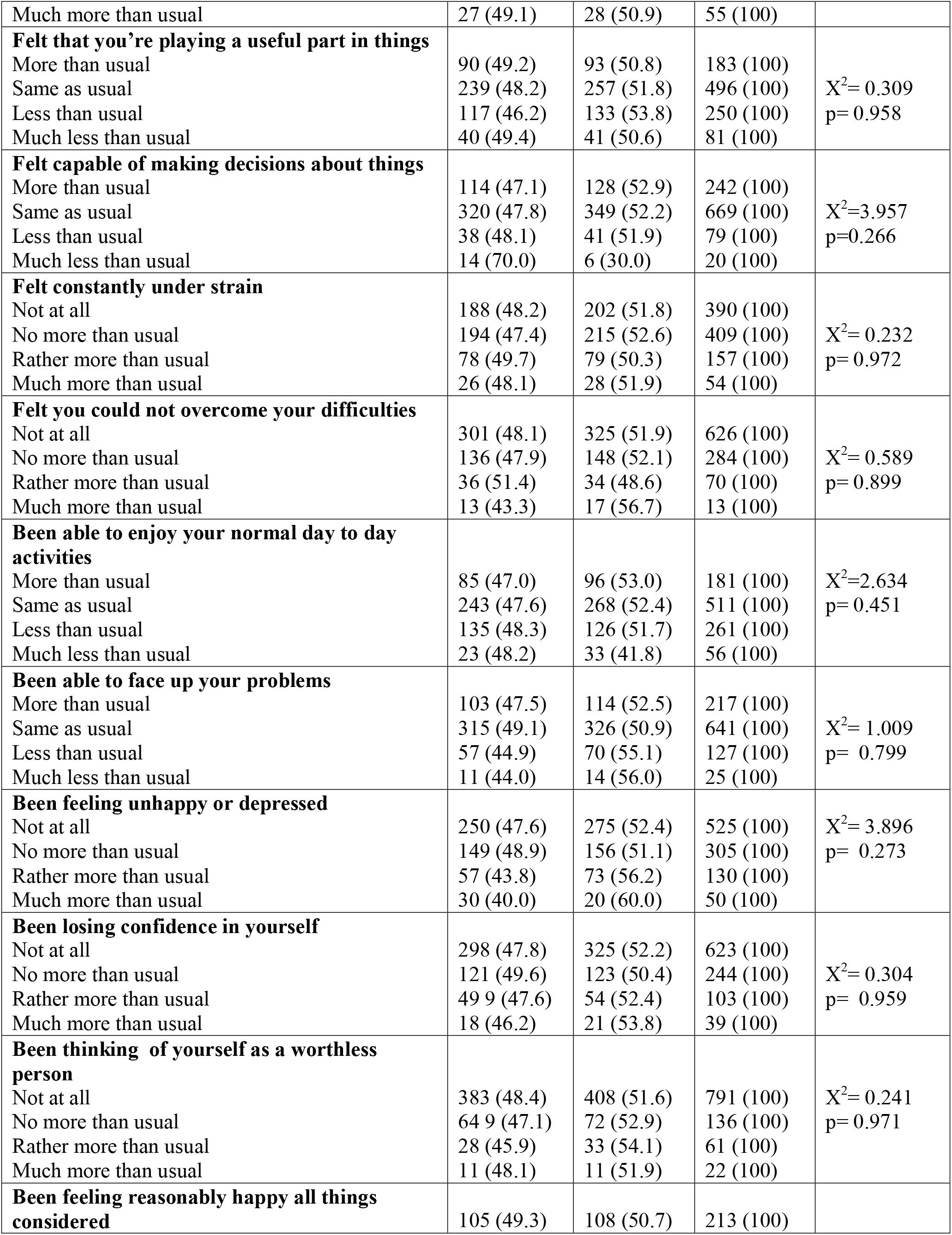
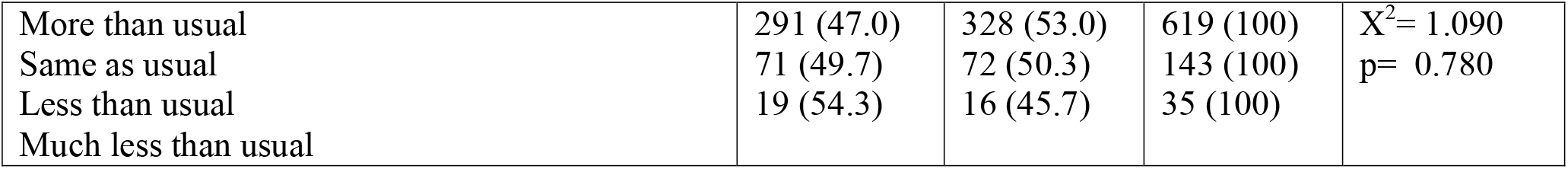
Responses of the participants to GHQ-12 Questions. *Have you during the COVID-19 pandemic*:

| Statement | Male<br>n (%) | Female<br>n (%) | Total | Chi-square<br>p value |
| --- | --- | --- | --- | --- |
| **Been able to concentrate on what you're doing** |  |  |  |  |
| Better than usual | 331 (48.4) | 353 (51.6) | 684 (100) | $X^2 = 0.339$<br>$p = 0.593$ |
| Same as usual | 119 (48.1) | 127 (51.6) | 246 (100) |  |
| Less than usual | 27 (45.0) | 33 (55.0) | 60 (100) |  |
| Much less than usual | 9 (45.0) | 11 (55.0) | 20 (100) |  |
| **Lost much sleep over worry** |  |  |  |  |
| Not at all | 79 (49.4) | 81 (50.6) | 160 (100) | $X^2 = 0.288$<br>$p = 0.962$ |
| No more than usual | 251 (48.3) | 269 (51.7) | 520 (100) |  |
| Rather more than usual | 129 (46.9) | 146 (53.1) | 275 (100) |  |
| Much more than usual | 27 (49.1) | 28 (50.9) | 55 (100) |  |
| **Felt that you're playing a useful part in things** |  |  |  |  |
| More than usual | 90 (49.2) | 93 (50.8) | 183 (100) | $X^2 = 0.309$<br>$p = 0.958$ |
| Same as usual | 239 (48.2) | 257 (51.8) | 496 (100) |  |
| Less than usual | 117 (46.2) | 133 (53.8) | 250 (100) |  |
| Much less than usual | 40 (49.4) | 41 (50.6) | 81 (100) |  |
| **Felt capable of making decisions about things** |  |  |  |  |
| More than usual | 114 (47.1) | 128 (52.9) | 242 (100) | $X^2 = 3.957$<br>$p = 0.266$ |
| Same as usual | 320 (47.8) | 349 (52.2) | 669 (100) |  |
| Less than usual | 38 (48.1) | 41 (51.9) | 79 (100) |  |
| Much less than usual | 14 (70.0) | 6 (30.0) | 20 (100) |  |
| **Felt constantly under strain** |  |  |  |  |
| Not at all | 188 (48.2) | 202 (51.8) | 390 (100) | $X^2 = 0.232$<br>$p = 0.972$ |
| No more than usual | 194 (47.4) | 215 (52.6) | 409 (100) |  |
| Rather more than usual | 78 (49.7) | 79 (50.3) | 157 (100) |  |
| Much more than usual | 26 (48.1) | 28 (51.9) | 54 (100) |  |
| **Felt you could not overcome your difficulties** |  |  |  |  |
| Not at all | 301 (48.1) | 325 (51.9) | 626 (100) | $X^2 = 0.589$<br>$p = 0.899$ |
| No more than usual | 136 (47.9) | 148 (52.1) | 284 (100) |  |
| Rather more than usual | 36 (51.4) | 34 (48.6) | 70 (100) |  |
| Much more than usual | 13 (43.3) | 17 (56.7) | 13 (100) |  |
| **Been able to enjoy your normal day to day activities** |  |  |  |  |
| More than usual | 85 (47.0) | 96 (53.0) | 181 (100) | $X^2 = 2.634$<br>$p = 0.451$ |
| Same as usual | 243 (47.6) | 268 (52.4) | 511 (100) |  |
| Less than usual | 135 (48.3) | 126 (51.7) | 261 (100) |  |
| Much less than usual | 23 (48.2) | 33 (41.8) | 56 (100) |  |
| **Been able to face up your problems** |  |  |  |  |
| More than usual | 103 (47.5) | 114 (52.5) | 217 (100) | $X^2 = 1.009$<br>$p = 0.799$ |
| Same as usual | 315 (49.1) | 326 (50.9) | 641 (100) |  |
| Less than usual | 57 (44.9) | 70 (55.1) | 127 (100) |  |
| Much less than usual | 11 (44.0) | 14 (56.0) | 25 (100) |  |
| **Been feeling unhappy or depressed** |  |  |  |  |
| Not at all | 250 (47.6) | 275 (52.4) | 525 (100) | $X^2 = 3.896$<br>$p = 0.273$ |
| No more than usual | 149 (48.9) | 156 (51.1) | 305 (100) |  |
| Rather more than usual | 57 (43.8) | 73 (56.2) | 130 (100) |  |
| Much more than usual | 30 (40.0) | 20 (60.0) | 50 (100) |  |
| **Been losing confidence in yourself** |  |  |  |  |
| Not at all | 298 (47.8) | 325 (52.2) | 623 (100) | $X^2 = 0.304$<br>$p = 0.959$ |
| No more than usual | 121 (49.6) | 123 (50.4) | 244 (100) |  |
| Rather more than usual | 49 9 (47.6) | 54 (52.4) | 103 (100) |  |
| Much more than usual | 18 (46.2) | 21 (53.8) | 39 (100) |  |
| **Been thinking of yourself as a worthless person** |  |  |  |  |
| Not at all | 383 (48.4) | 408 (51.6) | 791 (100) | $X^2 = 0.241$<br>$p = 0.971$ |
| No more than usual | 64 9 (47.1) | 72 (52.9) | 136 (100) |  |
| Rather more than usual | 28 (45.9) | 33 (54.1) | 61 (100) |  |
| Much more than usual | 11 (48.1) | 11 (51.9) | 22 (100) |  |
| **Been feeling reasonably happy all things considered** |  |  |  |  |
|  | 105 (49.3) | 108 (50.7) | 213 (100) |  |
| More than usual | 291 (47.0) | 328 (53.0) | 619 (100) | $X^2 = 1.090$<br>$p = 0.780$ |
| Same as usual | 71 (49.7) | 72 (50.3) | 143 (100) |  |
| Less than usual | 19 (54.3) | 16 (45.7) | 35 (100) |  |
| Much less than usual |  |  |  |  |

### Risk of psychological distress

Results from Table 3 showed that almost a third (31.4%) of the respondents were at risk of psychological distress and females constituted majority(60.3 %)as compared to their Male counterparts (39.7%) this was statistically significant (p < 0.001)

**Table 3:** Respondents showing risk of Psychological distress.

| Risk of Psychological distress | Male n (%) | Female n (%) | Total | Chi-square<br>p value |
| --- | --- | --- | --- | --- |
| No risk | 360 (58.9) | 333 (8.9) | 693 (100) | $X^2 = 12.969$<br>$p = <0.001$ |
| At risk | 126 (39.7) | 191 (60.3) | 317 (100) |  |
| Total | 486 (48.1) | 524 (51.9) | 1010 (100) |  |

### Predictors of psychological distress among all respondents

From Table 4; *Age* (OR=0.929, p=0.016), *female gender* (OR=1.534, 95% p=0.003), *family monthly income greater than 50,000 naira* (OR=1.378, p=0.023), and positive *history of mental illness* (OR=3.077, p=0.004) were independently associated with the risk of psychological distress among all respondents in general.

**Table 4:**
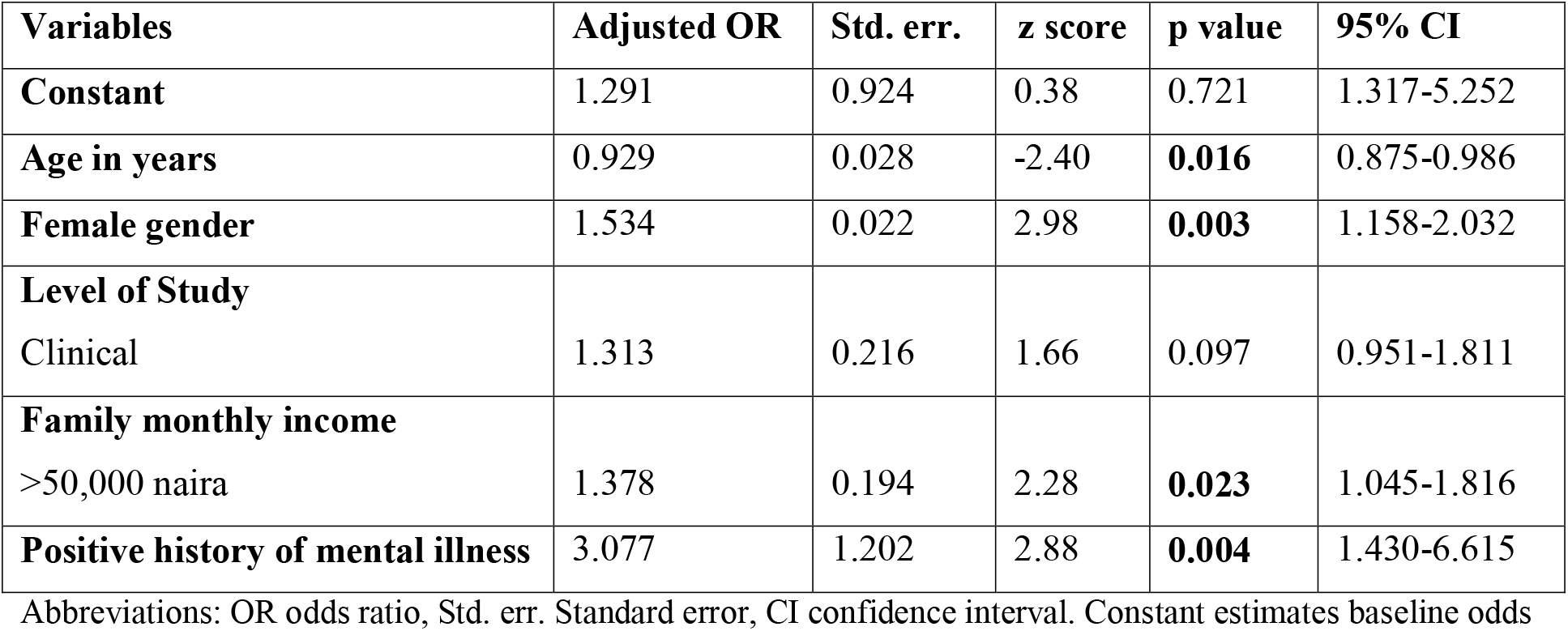
Socio-demographics and predictors of psychological distress among all respondents.

### Predictors of psychological distress among female respondents

Table 5 shows that *age* (OR=0.896, p=0.025), *family income greater than #50,000* (OR=0.530, p=0.001) and positive history of mental illness (OR=5.102, p=0.002) were independently associated with the risk of psychological distress among female participants.

**Table 5:** Socio-demographics and predictors of psychological distress among female respondents.

| Variables | Adjusted OR | Std. err. | z score | p value | 95% CI |
| --- | --- | --- | --- | --- | --- |
| **Constant** | 7.197 | 7.996 | 1.78 | 0.076 | 0.816-63.499 |
| **Age in years** | 0.896 | 0.044 | -2.25 | **0.025** | 0.814-0.986 |
| **Level of Study** |  |  |  |  |  |
| Clinical | 1.156 | 0.271 | 0.62 | 0.535 | 0.731-1.829 |
| **Family monthly income** |  |  |  |  |  |
| >50,000 naira | 0.530 | 0.101 | -3.35 | **0.001** | 0.366-0.769 |
| **Positive history of mental illness** | 5.102 | 2.647 | 3.14 | **0.002** | 1.845-14.106 |

Abbreviations: OR odds ratio, Std. err. Standard error, CI confidence interval. Constant estimates baseline odds

### Predictors of psychological distress among male respondents

From Table 6, there was no socio-demographic factor independently associated with the risk of psychological effects among male participants.

**Table 6:**
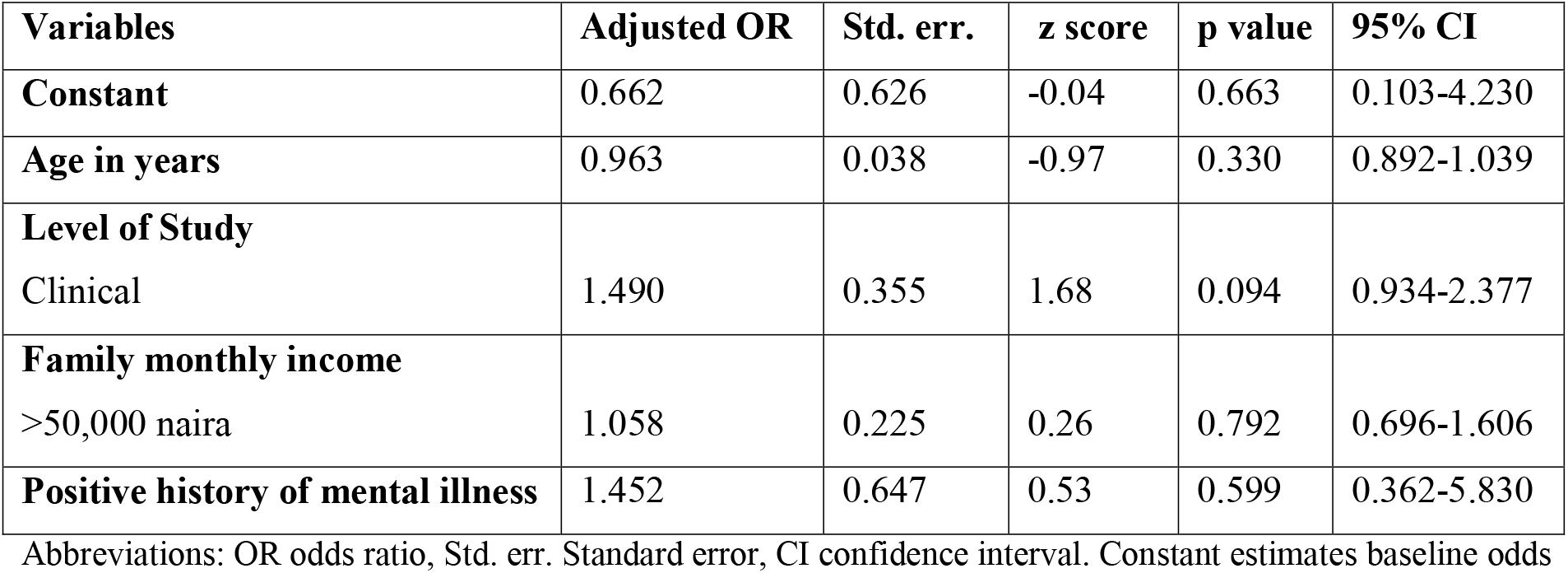
Socio-demographics and predictors of psychological distress among male respondents.

| Variables | Adjusted OR | Std. err. | z score | p value | 95% CI |
| --- | --- | --- | --- | --- | --- |
| Constant | 0.662 | 0.626 | -0.04 | 0.663 | 0.103-4.230 |
| Age in years | 0.963 | 0.038 | -0.97 | 0.330 | 0.892-1.039 |
| Level of Study<br>Clinical | 1.490 | 0.355 | 1.68 | 0.094 | 0.934-2.377 |
| Family monthly income<br>>50,000 naira | 1.058 | 0.225 | 0.26 | 0.792 | 0.696-1.606 |
| Positive history of mental illness | 1.452 | 0.647 | 0.53 | 0.599 | 0.362-5.830 |

Abbreviations: OR odds ratio, Std. err. Standard error, CI confidence interval. Constant estimates baseline odds

Female respondents had higher risk of psychological distress compared to males. However, the predictors of psychological distress among the female participants are age, family income greater than 50,000 naira and a positive history of mental health condition. Whereas, there was no predictor of psychological distress among male respondents.

## Discussion

The findings our study showed that the pandemic had a negative psychological impact on the respondents and agrees with Gupta et al, which stated that the negative psychological impact on the participants was greater during the lockdown due to COVID-19 pandemic [22]. From their survey, it was reported that the pandemic and lockdown increased mental morbidity among the general public [22] and the number of increasing cases resulted in an extension of the lockdown and curfews [22]. However, the results from our study and that of Gupta et al, contradict the claims of Yun Li et al [23] where all participants had low mean scores for anxiety and depression. This reason for this unusual finding may be because, as at the time of study, the number of COVID-19 cases was fewer compared to the latter period.

Our study reports that there is a significant gender difference in the psychological distress on medical students with females constituting a majority (60%) of the population at risk of psychological distress, this finding agrees with existing evidence from Ochilbek et al, which stated that females were at a higher risk of psychological distress than males [24]. In the study by Ochilbek and Senol, there was a significant gender difference as women had higher anxiety and depressionscores than men which confirm our results. The findings from our study further supports a study by Rodríguez-Rey et al which also revealed women showed significantly higher levels in anxiety, stress and depression [25]. In addition, a similarsurvey by Li G et al found out that female healthcare workers were more exposed to the psychological threat of COVID-19 than male healthcare workers [26]. However, the significant gender difference in this study is in contrast with findings from a similar pre-pandemic study which showed gender not to be significant in the psychological impact on the students. This may be because their study was done in the absence of a large scale stressor such as the pandemic [27].

Our results from the regression analysis indicated that female participants with family income greater than #50,000 are at risk of psychological distress compared to femaleparticipants who had lower family income. This is consistent with the study by Tull et al which discovered that income level was uniquely inversely associated with anxiety, financial worry and loneliness during the COVID-19 pandemic [28]. Our study also agrees with Saurabh et al which found out that the psychological problems of children and adolescents was mainly associated with loss of father’s job, financial losses of family and unavailability of basic life needs [29].

Furthermore, our regression analysis showed that female respondents who have been previously diagnosed with a mental disorder are also at increased risk of psychological distress, compared to female respondents who have never been diagnosed of a mental health condition. This finding further buttresses the theory that there is an association between mental disorders and stressful events.

This study is among the first few gender comparative studies in Nigeria that assessed the psychological distress of the COVID-19 pandemic on medical students, it also featuresa large sample size of 1010 distressamong Nigerian medical students. However it has some limitations and the findings should be interpreted with some caution.

Information obtained was collected using an online self-administered questionnaire indicating the possibility of selection bias. Furthermore, the results cannot be generalized to the entirecountry because it was limited to participants in three universities only in the South West region of Nigeria. There is also a possibility for recall bias and causal inferences cannot be made since due to the cross-sectional nature of the study

A more representative sample and a broader study involving colleges of medicine across the entire country would provide more generalized and accurate estimate of the gender comparison of the psychological distress of COVID-19 in medical students.

## Conclusion

Females were at higher risk of psychological distress compared to male students emphasizing the need for gender specific interventions for psychological distress among medical students. Also, future research to establish the long term psychological effects of the COVID-19 pandemic on medical students is recommended.

## Data Availability

All data referred to in the manuscript have been made available in the body of the manuscript.

## Acknowledgement

We appreciate Ademuyiwa EA, Bankole OM, Oaikhina OI, Dairo OO, Adetoye TA, Oyenuga OO, Owate OO, Kadiri NA and Oloyede OV, who assisted in data collection across the three colleges. Adesanya OA, Anibaba OM, Fashina IO, for providing technical help and general support, and Dr. Ogunyemi AO, for providing writing assistance.

